# Systematic review and meta-analysis of evidence on the efficacy of e-cigarette use for sustained smoking and nicotine cessation

**DOI:** 10.1101/2020.11.02.20224212

**Authors:** E Banks, A Yazidjoglou, S Brown, L Ford, T Zulfiqar, O Baenziger, G. Joshy

## Abstract

**Objective:** To systematically review and meta-analyse evidence regarding the efficacy of electronic nicotine delivery systems (ENDS) as smoking cessation aids.

**Data Sources:** PubMed, Scopus, Web of Science, PsycINFO, MEDLINE and Cochrane Library were searched up to February-March 2020 (PROSPERO registration CRD42020170692).

**Study selection:** Published peer-reviewed randomised controlled trials (RCTs) of the efficacy of ENDS for sustained cessation of combustible tobacco smoking and/or nicotine use, compared with no intervention, placebo or nicotine replacement therapy (NRT) by intention-to-treat, with a minimum of four months follow-up.

**Data Extraction:** Data were extracted independently into a pre-specified template. Risk of bias was assessed with the Cochrane Collaboration’s tool and evidence quality rated using GRADE.

**Data Synthesis:** From 3,973 titles identified, nine RCTs were identified; 330 of 5,445 smokers randomised quit. Smoking cessation did notPublic health consequences differ significantly for randomisation to ENDS versus: no intervention (three studies, random-effects meta-analysis RR 1.95; 95%CI 0.90-4.22); placebo (three studies, 1.61; 0.93-2.78) or NRT (three studies; 1.25; 0.74-2.11). Fixed-effects sensitivity analyses showed significant results for ENDS vs NRT (1.43; 1.10-1.86). Smokers randomised to ENDS were substantially more likely than control to use nicotine at follow-up. Overall evidence quality was low. Considering only studies without potential competing interests further limited evidence but did not materially change conclusions.

**Conclusions:** There is insufficient evidence that ENDS are efficacious for smoking cessation compared to no intervention, placebo or NRT. Results are promising, particularly for therapeutic use, but vary according to analytic method. ENDS may lead to greater ongoing nicotine exposure than other smoking cessation methods.

**Implications:** This systematic review and meta-analysis consolidates current evidence on the efficacy of ENDS as an aid to sustained smoking and nicotine cessation and considers these findings in the context of potential competing interests. While findings are promising more research - preferably independent of industry funding - is needed to provide robust evidence of the efficacy of ENDS for smoking cessation. Future research should investigate nicotine cessation in addition to smoking cessation.

## Background

The serious health risks of combustible tobacco smoking are well established. In 2019, The World Health Organization reported that tobacco smoking is responsible for over eight million deaths globally each year.^1^ The prevalence of smoking is falling in many countries as the result of reduced smoking initiation in younger generations and quitting among established smokers.^2^ Smoking cessation results in numerous health benefits, including dramatically reducing the risk of premature mortality, cardiovascular disease and cancer, compared with continuing to smoke. Although the majority of people who quit smoking successfully do so unaided,^3-8^ a range of regulator-approved smoking cessation medications are available, including nicotine replacement therapies (NRTs) such as chewing gums, nasal sprays, inhalers and transdermal patches. NRTs are safe, inexpensive, easily accessible and increase the likelihood of quitting, however their efficacy declines with long-term use and smoking relapse is common.^9^

Electronic nicotine delivery systems (ENDS) or e-cigarettes are often marketed – explicitly or implicitly – as effective and safe tools for smoking cessation, and smoking cessation is a common justification for use.^10^ These are devices containing heating units which vaporise a liquid, typically described as an ‘e-liquid’. E-liquids vary widely, but they generally contain propylene glycol, flavourings and nicotine, although electronic non-nicotine delivery systems (ENNDS) are also available.^11^ Nicotine is the main addictive substance in combustible cigarettes and ENDS. Concerns have been raised about the direct health effects of nicotine, particularly for children and youth.^12^

In order to maximise population health, policy and practice for e-cigarettes needs to draw on contemporary evidence that is of sufficient scale and quality to be reliable. The evidence on e-cigarettes and smoking cessation is evolving rapidly and repeated quantitative reviews of studies are necessary to ensure informed policy and practice.

Over recent years, major reports and published reviews have found that the current evidence is insufficient to conclude that e-cigarettes are effective as an aid to smoking cessation, compared with no treatment, placebo or other NRT.^13-18^ While observational data can provide important evidence on patterns of use of e-cigarettes and on certain health impacts, randomised controlled trials (RCTs) are the only study type able to consistently provide reliable evidence on the efficacy of interventions on their target endpoints, including evidence regarding e-cigarettes and smoking cessation. Many earlier reports and reviews have included observational data, as well as data from RCTs and all but the most recent of these reviews^19-21^ were only able to include evidence from three relatively small RCTs.^22-24^ No reviews, to our knowledge, have quantitatively considered the impact of potential competing interests and the impact of e-cigarettes on cessation of ongoing exposure to nicotine has not been reviewed to date.

This systematic review and meta-analysis aims to summarise the current published peer-reviewed RCT evidence on the efficacy of e-cigarettes – with or without nicotine – for the sustained cessation of combustible tobacco cigarette smoking and for the cessation of ongoing exposure to nicotine. The review also considers the evidence in the light of potential competing interests.

## Methods

### Search Strategy

A systematic review was undertaken to examine the efficacy of e-cigarettes as a smoking cessation aid and was consistent with that used in a recent national US report.^13^ Six databases (PubMed, Scopus, Web of Science, PsycINFO (Ovid), MEDLINE (Ovid), and Cochrane Library) were searched between 5 February and 2 March 2020. There was no date limit. The systematic review protocol was published on PROSPERO (CRD42020170692). Further details on search terms are located in Supplementary material Appendix 1.

### Inclusion and Exclusion Criteria

This review included RCTs, as defined by the Cochrane Community,^25^ in which current smokers of any age were randomised to e-cigarettes or to no e-cigarettes, other smoking cessation treatment interventions (e.g. approved NRTs, behavioural therapy, combination), or placebo. The outcomes included were biochemically verified sustained cessation of combustible tobacco smoking and, separately, nicotine cessation (i.e. cessation of combustible tobacco smoking, ENDS or other NRT). Studies with cessation outcomes measured earlier than four months after their quit date were excluded in accordance with standard measures of sustained abstinence and outcomes at the latest follow-up date were included.^13,16,17^ All other study designs or populations were excluded. For further details refer to the Supplementary material Appendix 1.

### Eligibility Screening

Papers were imported into an EndNote library, exported to Covidence^26^ and duplicates were removed. Two authors independently screened all titles, abstracts and full-texts based on the inclusion and exclusion criteria. Only studies with abstracts published in English were screened. Forward and backward reference searching was performed from the final included articles using the ANU Library, Web of Science and Scopus. Discrepancies were resolved through discussion with a third author.

### Data Extraction

Two authors independently extracted the relevant data using a pre-specified data extraction template. Relative risks and 95% confidence intervals – by intention to treat – were extracted from each paper. In studies where relative risks were not presented, where possible, these were calculated from the number of events or percentages reported in the published study. Further details are in Appendix 1 of the Supplementary material.

Research funding and author conflict of interest information was extracted from each study. Studies were grouped based on whether or not authors or the study reported receiving funding and/or contributions in kind for the study from the tobacco or e-cigarette industry.

### Statistical Analysis

Where appropriate, relative risks from studies were combined using meta-analyses to assess the efficacy of ENDS for smoking cessation compared to no intervention (or usual care), placebo (ENNDS) and to NRT. Following data extraction but prior to any meta-analyses we assessed whether random- or fixed-effect models were most appropriate. Due to the likelihood that the interventions and the target populations in the different studies differed materially, a random-effects REML model was used for the primary analyses. The I-squared statistic was used to evaluate statistical heterogeneity between studies. Because the small number of studies for each outcome made random-effects modelling less suitable, we conducted sensitivity analyses using fixed-effects modelling. Other sensitivity analyses included repeating the analyses restricted to studies without noted potential competing interests and, separately, examining outcomes at the most consistent sustained follow-up time available (i.e. 24-26 weeks). All analyses were conducted using STATA version 16.1.

### Quality Assessment

The risk of bias for each included RCT was assessed independently by two authors using the Cochrane Collaboration’s tool for assessing risk of bias in randomised trials.^27^ The quality of the body of evidence for smoking cessation was evaluated using the GRADE approach.^28^ No studies were excluded based on their quality assessment scores.

## Findings

### Literature Search Result

Of the 3,973 titles identified for screening, nine eligible RCTs of ENDS were identified that examined smoking cessation as an outcome and were included in this review (Figure 1). There were no RCTs which examined the efficacy of ENNDS for smoking cessation as the primary outcome or that examined nicotine cessation as their primary outcome. The characteristics of the eligible studies are presented in Table 1 and Table S1 of the Supplementary material. A total of 5,445 smokers were randomised in studies conducted from 2013-2020; 330 achieved smoking cessation at follow-up: 2,836 randomised to ENDS and 2,609 to comparison groups.

**Table 1:**
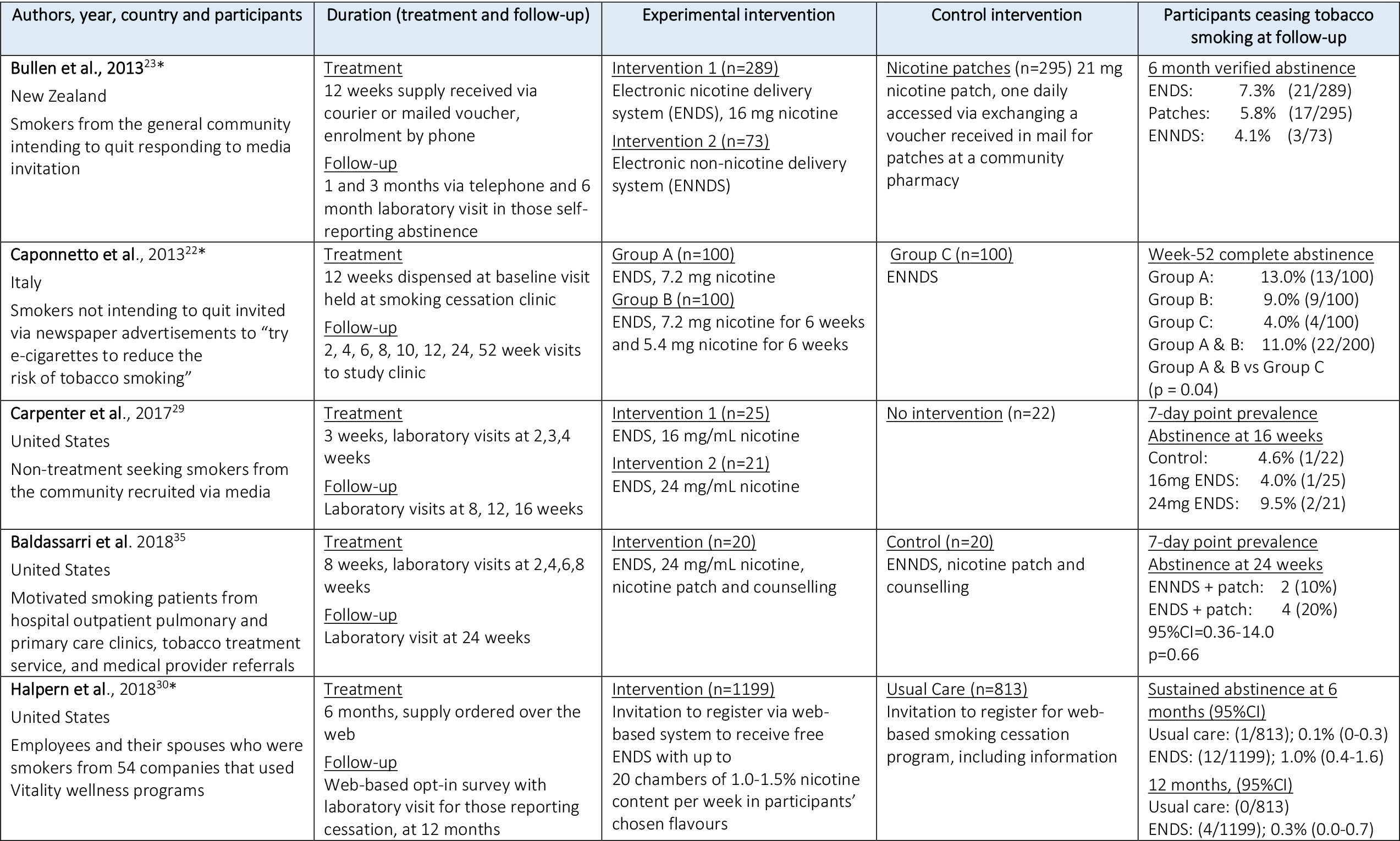
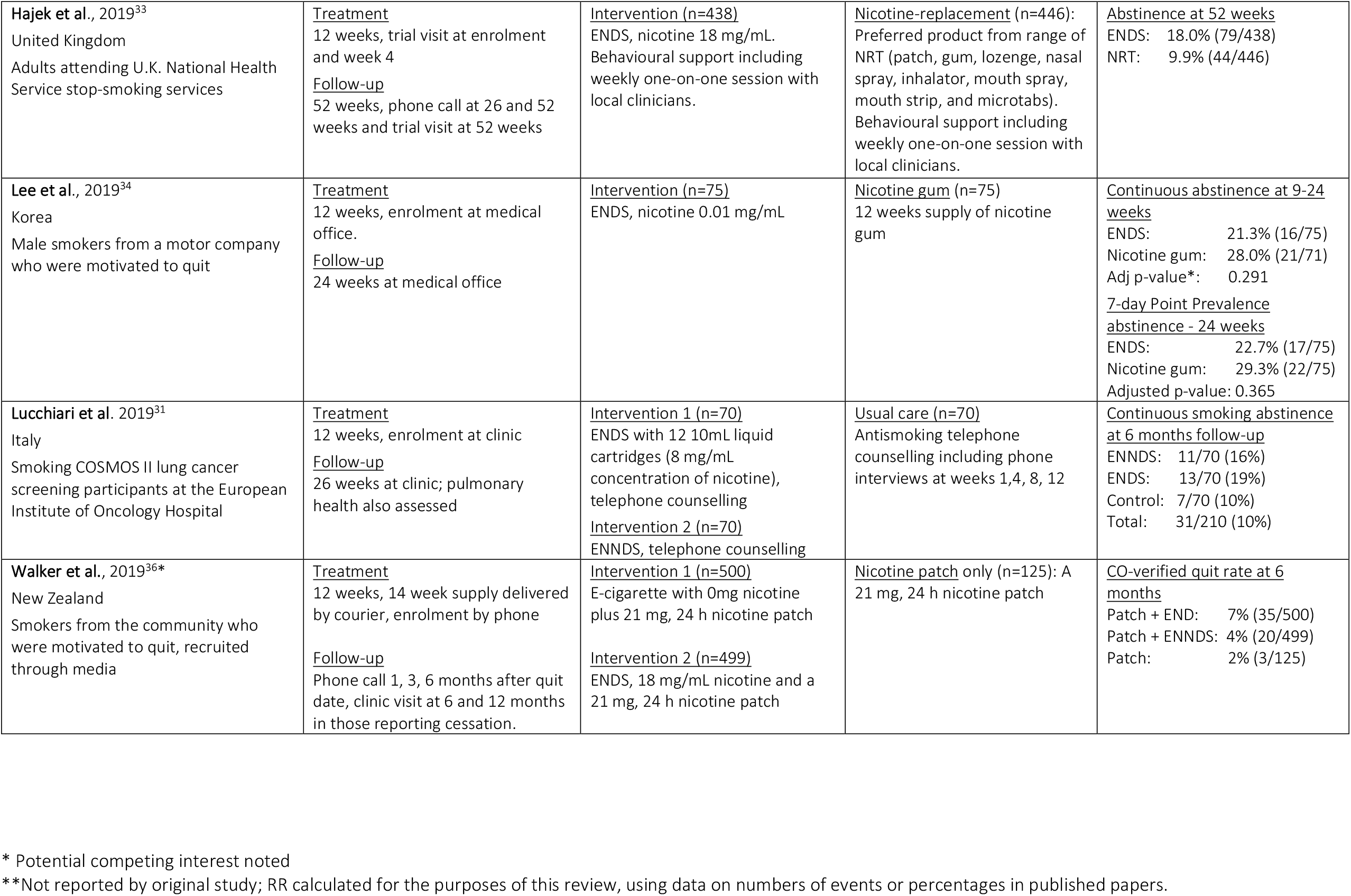
Details from identified RCTs of nicotine-delivering electronic cigarettes with smoking cessation as a primary outcome.

**Figure 1:**
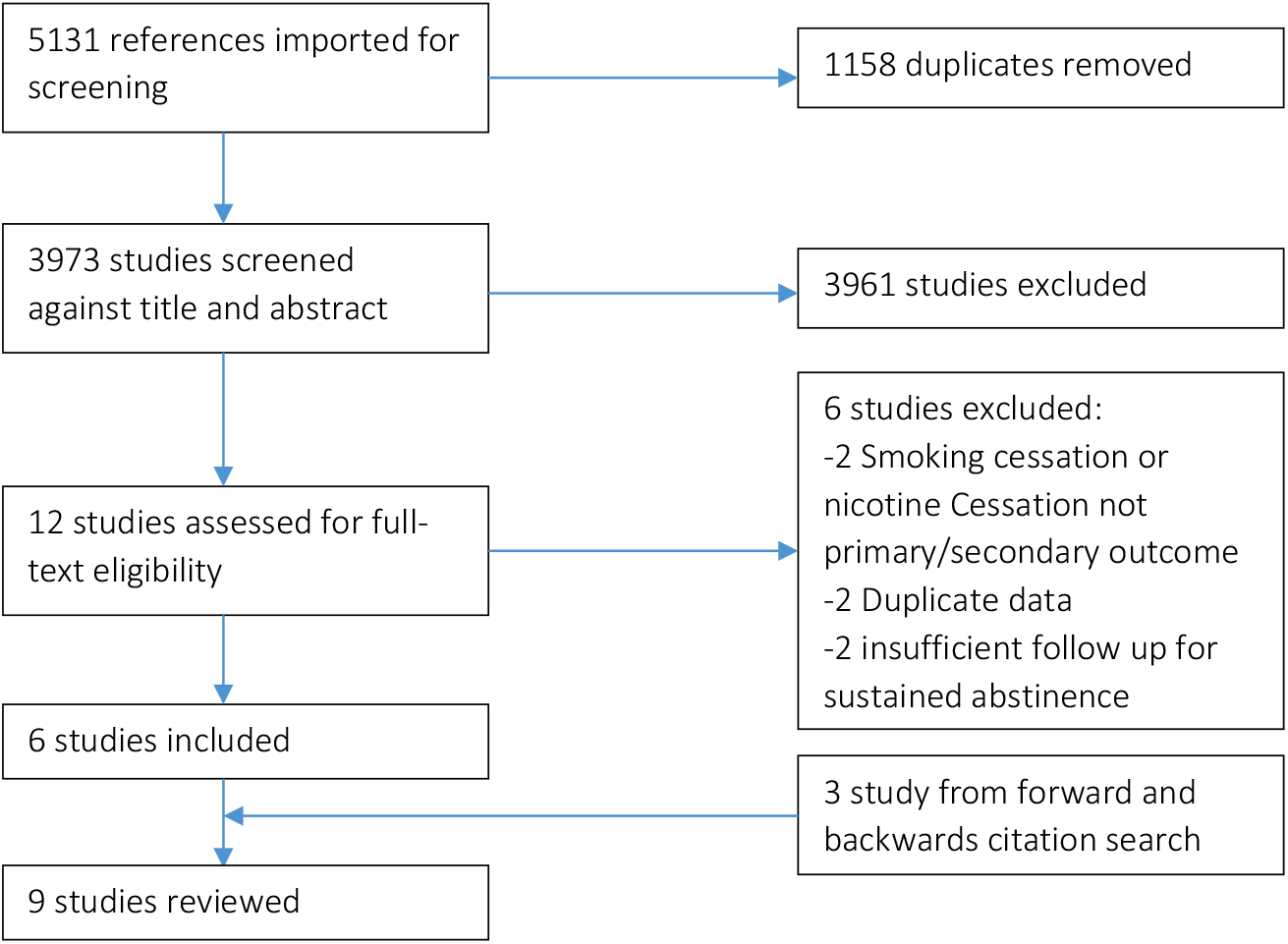
E-cigarette and smoking cessation review flowchart.

### Nicotine-delivering e-cigarettes versus no intervention or usual care

Three RCTs compared ENDS to no intervention or usual care (Table 1 and Table S1).^29-31^ These studies randomised a total of 2,220 participants, of whom 28 achieved sustained smoking cessation (Figure 2). None were funded directly by the tobacco or e-cigarette industry, nor were there any reported potential competing interests for the authors of the studies. Halpern et al reported receiving e-cigarettes donated by an e-cigarette company.^30^

**Figure 2:**
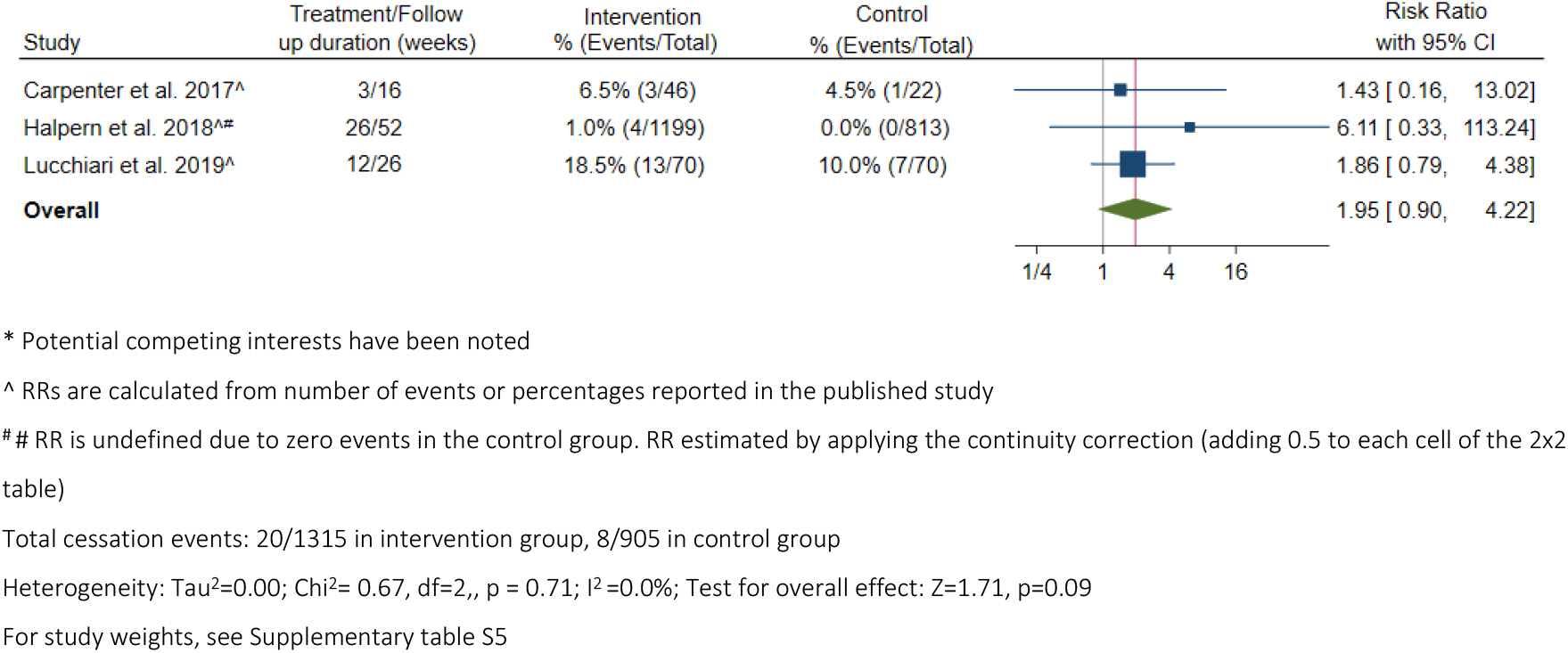
Biochemically verified sustained smoking cessation in smokers randomised to nicotine-delivering e-cigarettes versus no intervention or usual care: random-effects meta-analysis.

In their pilot RCT, Carpenter et al recruited 68 community-dwelling US smokers via media outlets who were not specifically seeking treatment.^29^ Participants were randomised to control or to three weeks of ENDS and attended multiple laboratory visits for follow-up. At four month follow-up, 4.0% of the 16mg and 9.5% of the 24mg nicotine ENDS groups versus 4.6% of the control (no intervention) respectively, achieved biochemically-verified 7-day point prevalent abstinence (RR ENDS versus control 1.43; 95% CI 0.16-13.02); this difference was not statistically significant.

Also in the US, the web-based RCT of Halpern et al included 6,006 smokers from employees and their spouses from companies that utilised Vitality wellness programs –2,012 in study arms comparing ENDS and usual care.^30^ Participants were contacted by email and accessed study interventions and reported outcomes via a web portal; no contact was assumed to represent continuing smoking and cessation outcomes were verified biochemically only in those reporting cessation. At six month follow-up, 12 of 1,199 participants (1.0%; 95% CI 0.4%-1.6%) in the ENDS arm and one of 813 participants in the usual care arm (0.1%; 95% CI 0.0%-0.3%) were verified as having ceased smoking. After accounting for multiple testing, there was no statistically significant difference in outcomes between these groups.^30^ At 12 month follow-up, 4 of 1,199 participants (0.3%; 95% CI 0.0%-0.7%) in the ENDS arm and none of 813 participants in the usual care arm were verified as having ceased smoking.

In a study recruiting smokers from an Italian screening program for lung cancer and including clinic-based follow-up and telephone smoking cessation counselling, Lucchiari et al found 19.0% of 70 smokers randomised to three months of ENDS and 10.0% of 70 smokers randomised to control achieved continuous biochemically verified abstinence at six month follow-up (RR 1.86; 95% CI 0.79- 4.38).^31^

No individual study reported a significant difference in cessation outcomes between randomised groups. Results from the random-effects meta-analysis also found no significant difference at 4-12 month follow-up (RR 1.95; 95%CI 0.90-4.22; I^2^ = 0.0%)(Figure 2) or at six month follow-up (RR 2.88; 95% CI 0.77-10.79) (Supplementary Figure S3). This conclusion did not change materially when a fixed-effects model was used (RR 2.08, 95%CI 0.96-4.48)(Supplementary table S5). Nor did it change substantively when the random-effects meta-analysis was restricted to studies with no noted potential competing interests (RR 1.80; 95%CI 0.81-3.99; I^2^ = 0.0%), although evidence was even more limited, with 24 of 208 participants ceasing smoking (Supplementary Figure S1). Two of the included studies were assessed as having a high risk of bias^29,30^ and one was found to have some concerns^31^ (Supplementary Table S3).

### Nicotine-delivering e-cigarettes versus e-cigarettes which do not deliver nicotine

Three RCTs compared smoking cessation outcomes in participants randomised to ENDS and ENNDS (considered a placebo) (Table 1 and Supplementary Table S1).^22,23,31^ These trials reported a total of 74 participants ceasing smoking out of 802 randomised (Figure 3). No studies were directly funded by the tobacco or e-cigarette industry. Bullen et al^23^ had a study author who reported previously receiving research funding from an e-cigarette manufacturer and Caponetto et al^43^ had a study author who had received funding from the tobacco industry.^32^ Both studies reported using e-cigarettes donated by an e-cigarette company.^22,23^

**Figure 3:**
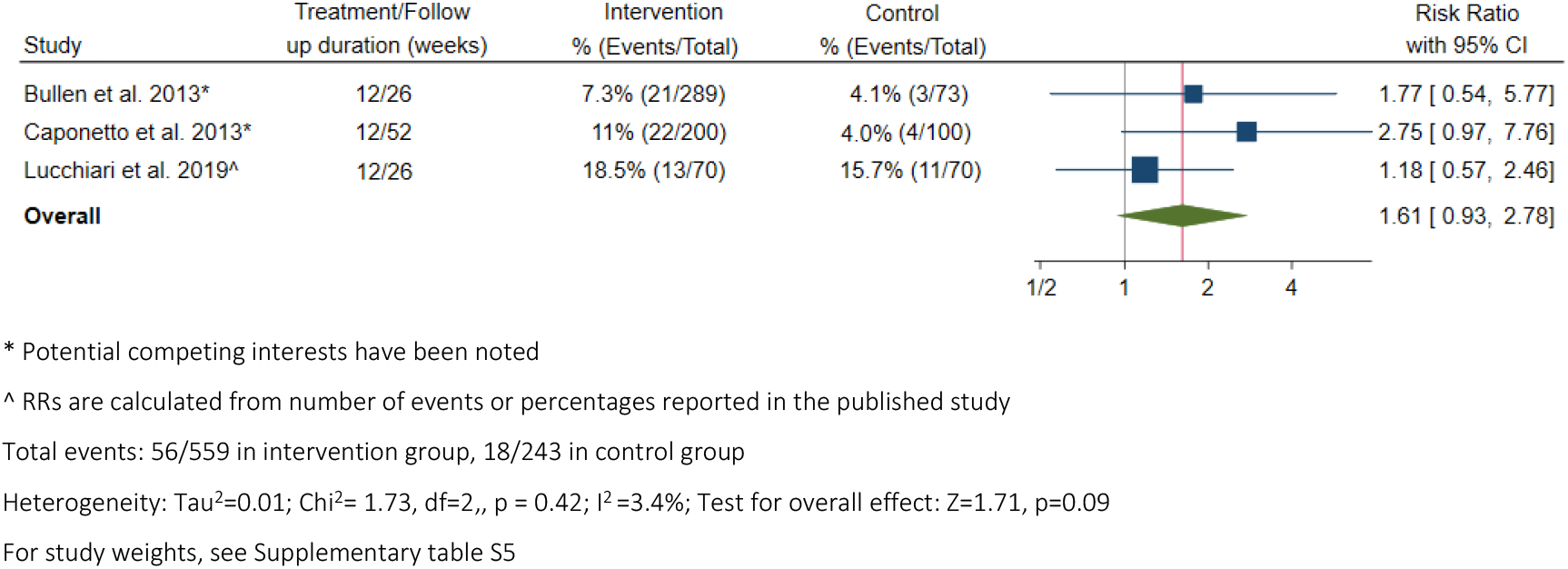
Biochemically verified sustained smoking cessation in smokers randomised to nicotine-delivering e-cigarettes versus non-nicotine-e-cigarettes: random-effects meta-analysis.

In their Italian pilot RCT published in 2013, Caponnetto et al recruited 300 smokers not intending to quit via newspaper advertisements inviting them to try e-cigarettes “to reduce the risk of tobacco smoking”.^22^ The study protocol included nine visits held at a smoking cessation clinic and participants received a 12 week supply of e-cigarettes at baseline. At one-year follow-up 11.0% (22/200) of participants randomised to ENDS and 4.0% (4/100) of participants randomised to ENNDS achieved cessation (RR 2.75; 95% CI 0.97-7.76).

In the New Zealand superiority RCT of Bullen et al, community-dwelling smokers who were motivated to quit were recruited through community newspapers. Participants telephoned a screening clinic and received interventions via courier; 289 were randomised to 12 weeks of 16mg nicotine e-cigarettes and 73 were randomised to 12 weeks of ENNDS. At six-month follow-up 7.3% (21/289) of smokers randomised to ENDS and 4.1% (3/73) randomised to ENNDS had verified smoking abstinence (RR 1.77; 95% CI 0.54-5.77).^23^

The Italian study of Lucchiari et al, outlined above, reported that 19.0% of smokers randomised to ENDS and 16.0% randomised to ENNDS achieved continuous abstinence at 6 month follow-up (RR 1.18; 95% CI 0.57-2.46).^31^

No statistically significant difference between ENDS and ENNDS was found in any study. The random-effects summary RR for smoking cessation at 6-12 month follow-up in those randomised to ENDS versus ENDS was 1.61, with no statistically significant difference between the groups (95%CI 0.93-2.78; I^2^=3.4%)(Figure 3). The finding was of borderline significance using fixed-effects meta-analysis (RR1.71, 95%CI 1.00-2.92)(Supplementary table S5) and did not change materially when restricted to six month follow-up only (RR 1.54; 95%CI 0.92-2.59) (Supplementary Figure S4). One of the included studies was assessed as having a high risk of bias^29^ and the remaining two were considered to raise “some concerns”^23,31^ (Supplementary Table S3). Restricting the evidence to that without known potential competing interests, one study remained with a RR of 1.18 (95%CI 0.57-2.46) for cessation in smokers randomised to ENDS versus ENNDS, based on 140 participants, 24 of whom quit successfully.^31^

### Nicotine-delivering e-cigarettes versus other nicotine replacement therapy

Three RCTs were identified that compared ENDS to other NRT (Table 1 and Supplementary Table S1).^23,33,34^ The studies were conducted between 2013 and 2019. They included a total of 1,618 participants, all of whom were smokers motivated to quit and were randomised to 12-week treatment programs; 198 achieved smoking cessation at greater than four-month follow-up. Bullen et al had the potential competing interests noted above; no other studies had reported competing interests.

In the previously mentioned New Zealand RCT, smoking cessation at six months was achieved by 7.3% (21/289) of those randomised to ENDS and 5.8% (17/295) of those randomised to nicotine patches (RR 1.26; 95% CI 0.68-2.34).^23^

In a study of patients attending the UK National Health Service smoking cessation program, Hajek et al randomised smokers to ENDS or to a range of NRT products as the comparator (patch, gum, lozenge, nasal spray, inhalator, mouth spray, mouth strip, and microtabs), encouraging participants in the NRT group to combine and/or switch products.^33^ Behavioural therapy was provided to all participants, including weekly one-on-one sessions with local clinicians for at least four weeks after the quit date. ^33^ This study found that 18.0% (79/438) of those randomised to ENDS and 9.9% (44/446) of those randomised to other NRT achieved one-year sustained abstinence from smoking (RR 1.83; 95% CI 1.30- 2.58).

Lee et al randomised male smoking employees at a motor company in Korea to either ENDS or nicotine gum; all participants received an education session and four-weekly visits to a medical office for evaluation and counselling by an independent medical practitioner.^34^ At 24-week follow-up, 21.3% (16/75) of the ENDS and 28.0% (21/75) of the nicotine gum groups achieved continued smoking abstinence (adjusted p=0.291; RR 0.76; 95% CI 0.43-1.34).

In summary, two studies reported no statistically significant difference between ENDS and other NRT^23,34^ and the other found significantly greater cessation in those randomised to ENDS^33^ (Figure 4). Results from the random-effects meta-analysis found that there was no statistically significant difference in the efficacy of ENDS compared to NRT for smoking cessation at 6-12 month follow-up, with substantial variation in these results (RR 1.25; 95% CI 0.74-2.11; I^2^ = 69.0%)(Figure 4). This finding was statistically significant using fixed-effects meta-analysis (RR 1.43; 95%CI 1.10-1.86) (Supplementary table S5). The conclusion from the random-effects model did not substantially change when the meta-analysis was limited to studies with no noted potential competing interests (RR 1.22; 95% CI 0.52-2.86; I^2^ = 85.1%), although evidence was even more limited, with 160 of 1,034 participants ceasing smoking (Supplementary Figure S2).The summary RR at six month follow-up was similar to that incorporating 12 month results (RR 1.18; 95% CI 0.82-1.70) (Supplementary Figure S5).

**Figure 4:**
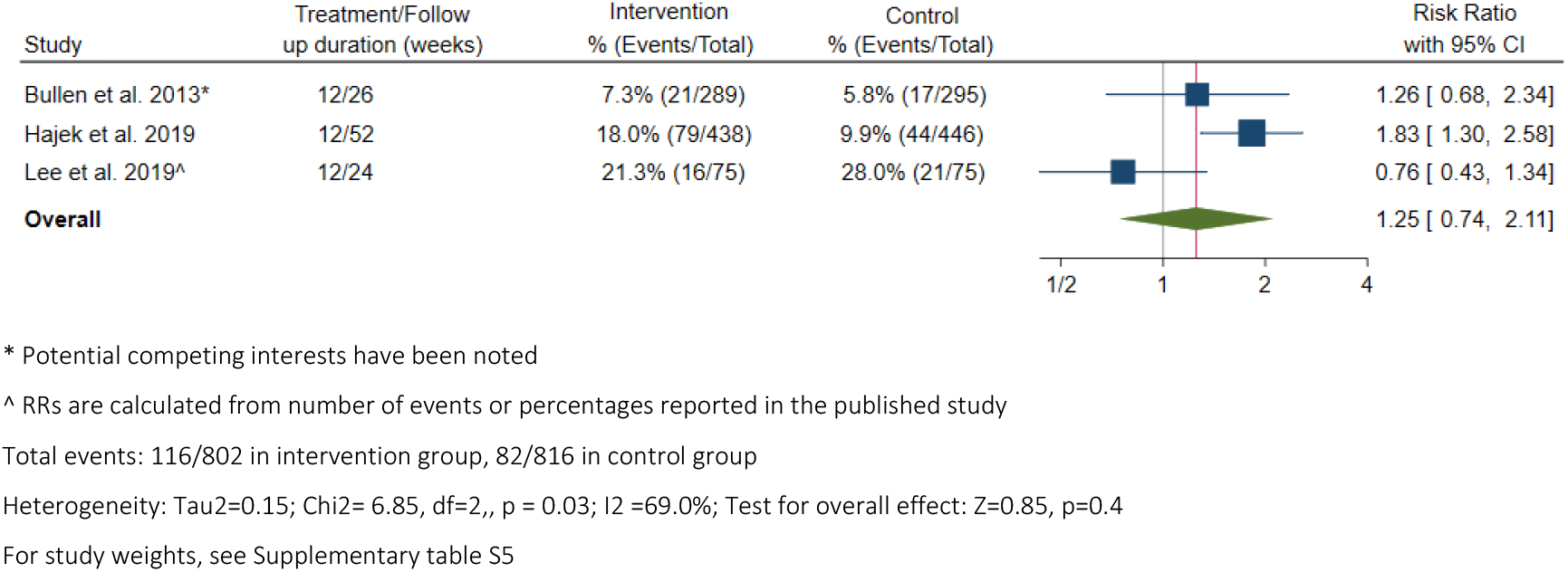
Biochemically verified sustained smoking cessation in smokers randomised to nicotine-delivering e-cigarettes versus other nicotine-replacement therapy: random-effects meta-analysis.

### E-cigarettes and other comparators

Two studies examined quitting in smokers randomised to ENDS and ENNDS, with all study participants receiving nicotine patches (Table 1 and Figure 5).^35,36^ One study had potential competing interests identified.^36^

**Figure 5:**
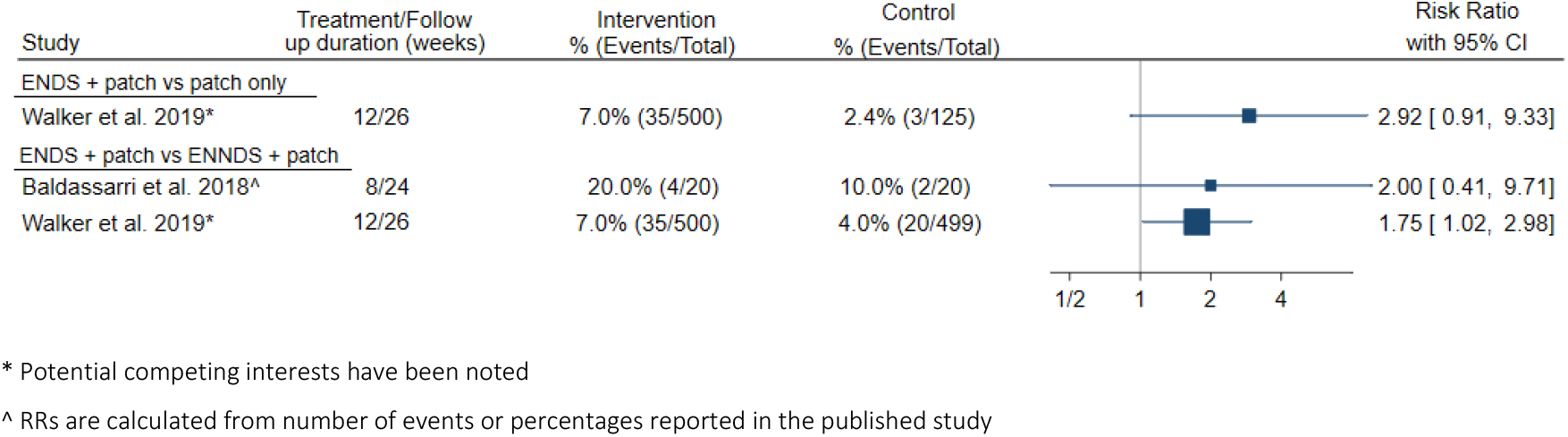
Biochemically verified smoking cessation in smokers using patches, randomised to nicotine-delivering e-cigarettes, non-nicotine e-cigarettes or no additional intervention.

In their US pilot RCT of 40 smokers willing to quit who were attending clinics and smoking cessation services, Baldassarri et al found that 20.0% randomised to ENDS and nicotine patches and 10.0% randomised to ENNDS and patches achieved 7-day point prevalence abstinence at 24 weeks (RR 2.00; 95% CI 0.41-9.71).^35^ Walker et al found that among New Zealand community-dwelling smokers, 7.0% (35/500) of motivated smokers randomised to 14 weeks of ENDS combined with nicotine patches achieved cessation at six months, compared to 2.4% (3/125) of those randomised to patches alone (RR 2.92; 95% CI 0.91-9.33)(Figure 5).^36^ Cessation was 4% (20/499) in smokers randomised to ENNDS plus nicotine patch (RR compared to patch only 1.75; 95% CI 1.02-2.98).

### Use of ENDS and nicotine cessation

There was limited evidence on the efficacy of ENDS as an aid to nicotine cessation, with no RCTs including this as an *a priori* outcome (Supplementary Table S2). Five RCTs contained data on nicotine cessation: two with^23,36^ and three without^29,33,35^ competing interests noted. These RCTs involved 2,773 smokers, 232 of whom quit during the follow-up period.

One study contained sufficient data to compare cessation of any nicotine exposure between participants randomised to ENDS or NRT.^33^ Data from Hajek et al indicate that 3.7% (16/438) of participants randomised to ENDS and 9.0% (40/446) of participants randomised to NRT had ceased all nicotine exposure (combustible cigarettes, ENDS or NRT) at 52 week follow-up (RR for ceasing any nicotine exposure=0.41; 95% CI 0.23-0.72).^33^

At 52 week follow-up in Hajek et al, 39.5% (173/438) of smokers randomised to ENDS were using nicotine-delivering products (ENDS or NRT) compared to 4.3% (19/446) of the NRT group, meaning smokers randomised to ENDS were 9.27 times (95% CI 5.88-14.61) as likely than those randomised to NRT to be using any nicotine-delivering products.^33^ Restricting the data to smokers who quit successfully, 79.8% (63/79) of quitters randomised to ENDS and 9.1% (4/44) of quitters in the NRT group were using nicotine-delivering products at 52 weeks (RR 8.77; 95% CI 3.42-22.48).^33^ Continuing smokers in the ENDS group were also much more likely to be using nicotine-delivering products at follow-up compared to those in the other NRT group (RR 8.21; 95% CI 4.88-13.82).^33^

In their New Zealand study published in 2013, Bullen et al.^23^ found that participants in the ENDS group were 4.26 times (95% CI 2.58-7.06) as likely to be using any nicotine-delivering products at six month follow-up compared with those randomised to other NRT. In the ENDS group, 38% (8/21) of combustible tobacco quitters were still using ENDS at follow-up. The number of participants still using other NRT in the other NRT group was not reported.

Data from the US pilot study conducted by Carpenter et al^29^ indicate that in the week preceding the final study visit (Week 16), 32.0% of participants in the 16mg ENDS group, 60.0% of participants in the 24mg ENDS group and 13.0% of participants in the control (no intervention) group were using ENDS.^29^

In the small Italian pilot study of Baldassarri et al^35^ at 24 week follow-up, 90.0% (18/20) of smokers randomised to ENDS and nicotine patch and 95.0% (19/20) randomised to ENNDS and nicotine patch were using nicotine in any form (combustible cigarettes, ENDS or other NRT)(RR for having ceased nicotine in any form for ENDS + patch versus ENNDS + patch 2.00; 95% CI 0.20-20.33).^35^ Among quitters, 50.0% (2/4) of the ENDS plus patch group and 50.0% (1/2) of the ENNDS plus patch were using NRT or e-cigarettes at follow-up (RR 1.00; 95% CI 0.18-5.46).^35^ Walker et al.^36^ found that intervention groups that included e-cigarettes were more likely to be using NRT products – including ENDS and other products – at six month follow-up, compared with the patch-only control group (ENDS + patch versus patch only RR 1.53; 95% CI 1.05-2.22; ENNDS + patch versus patch only RR 1.52; 95% CI 1.05-2.21).

In summary, the evidence regarding e-cigarette use in smokers and nicotine cessation is very limited. Considering the data that are available, smokers using e-cigarettes are substantially more likely to be using nicotine in any form (combustible cigarettes, ENDS or other NRT) at six to 12 month follow-up, or to be using ENDS or other NRT, than smokers who used other forms of NRT. There were insufficient data to compare ENDS and no intervention. Restricting data to studies without potential competing interests had no material effect on the conclusions.

### Quality assessment

Six of the nine studies were found to have a high risk of bias^22,29,30,34-36^ two raised some concerns^23,31^ and one was found to have a low risk of bias^33^ (Supplementary Table S3). Risk of bias did not appear to vary according to whether or not the study had noted potential competing interests. The overall quality of the evidence using GRADE was rated as low (Supplementary Table S4).

## Discussion

The results from this systematic review indicate that it is unclear whether e-cigarettes – nicotine-delivering or without nicotine – are efficacious as aids for smoking cessation. Large-scale reliable independent evidence is lacking; with only nine relevant RCTs identified, most of which are small and have methodological issues; the overall quality of the evidence is rated as low. Random effects meta-analyses based on these limited data do not show any significant benefit of nicotine-delivering e-cigarettes for smoking cessation, compared to no intervention/usual care, ENNDS or approved NRT.

Results using a different analytic method showed similar results for ENDS versus no intervention/usual care or ENNDS but significantly greater cessation in smokers to randomised to ENDS versus approved NRT, largely driven by the results of a single trial in UK therapeutic smoking cessation services. Hence, the evidence is not robust but is promising that ENDS may help with cessation, supporting the need for additional high-quality large-scale RCTs.

Studies of NRT receiving funding from industry, and sponsored device and drug studies more broadly, tend to find more favourable results than those without such funding.^37,38^ When the review and meta-analyses were restricted to studies with no apparent potential competing interests, evidence on e-cigarettes and smoking cessation became even more limited, although the general direction of the findings did not change materially.

ENDS deliver nicotine, so it is plausible that they would support cessation in ways similar to other products that deliver nicotine. It has been proposed that e-cigarettes may have advantages over other NRTs. They involve certain behavioural and sensory aspects of smoking, such as hand-mouth movement,^39^ and can rapidly and directly deliver nicotine to the user at relatively high doses. Hence, they have greater similarity to the combustible cigarette experience, which may increase efficacy for cessation, as well as the risk of abuse and long-term use.^40-43^

At the same time, use of ENDS may potentially support continuing smoking and dual use of combustible tobacco cigarettes and e-cigarettes is one of the most common patterns of observed use.^44-46^ High cost, limitations on places where smoking is allowed, bans on advertising, clear health warnings and reduced social acceptability are all important elements in comprehensive tobacco control.^47^ Smokers may potentially be able to mitigate some of these impacts through dual use with ENDS, thereby prolonging smoking, as ENDS are generally cheaper than smoking, are often able to be used in settings where combustible cigarettes are prohibited, their health impacts are less clear and they are often more socially acceptable.

If ENDS are used as a tobacco cessation tool, and use continues following cessation, there is ongoing exposure to nicotine, a highly addictive drug.^39,48,49^ There are concerns that nicotine addiction itself is problematic and that, although ENDS use would generally be considered better than continuing to smoke, quitting nicotine altogether is preferable. The use of nicotine-delivering e-cigarettes tends to result in more prolonged exposure to nicotine than use of other types of NRT. In an RCT based in the UK National Health Service, almost 80% of combustible tobacco smoking quitters randomised to ENDS were still using them one year following their quit date, and were almost nine times more likely to be using any nicotine-delivering product at follow-up compared to quitters in the NRT arm.^33^ Findings were similar in participants who continued to smoke.^33^ A letter to the editor about this RCT notes, “For every 100 participants who used the e-cigarette strategy, 18 quit smoking, but 14 of those participants became e-cigarette users. An additional 25 participants who did not quit smoking became dual users, so the e-cigarette strategy created more dual users than quitters, and most participants who quit smoking transitioned to vaping”.^50^

The available evidence on e-cigarettes and smoking cessation is affected by significant methodological issues. Many of the trials are small, with three explicitly termed pilot studies, designed more to test future study feasibility than the efficacy of e-cigarettes for cessation. The overall number of smokers quitting is also small: 197 in those randomised to ENDS and 133 in those randomised to comparators. This contributes to the lack of statistical power for the body of evidence as a whole to both detect and exclude an effect. It also makes publication and other types of bias more probable, including the fact that researchers may be more likely to choose not to publish negative findings from small studies.^51^ The small number of relevant RCTs means tests for funnel plot asymmetry is not appropriate to investigate the potential for publication bias.^52^ Loss to follow up and issues with ascertainment of cessation are also issues, especially for trials involving minimal contact with participants. The RCT including the largest number of participants, randomising employees at multiple US companies, recorded that none of the 813 smokers in the control arm had quit over a twelve month period. As well as being relatively statistically unstable, this is not consistent with the background 12-month quit rate in the general US population.^53^ In this web-based study, participants needed to actively log on to record smoking outcomes – no activity was taken to indicate continuing smoking – as well as to access intervention e-cigarettes. It is therefore likely that cessation events were missed in study participants and possible that those in the ENDS intervention arm had greater engagement and reporting of outcomes.

The generalisability of the RCT evidence is also problematic. E-cigarettes are highly heterogeneous, with many thousands of variants in the e-liquid used, including the dose and nature of the nicotine delivered. There was major variation in the settings and participants of the included RCTs, ranging from minimal contact telephone- and web-based studies of smokers with or without specific plans to quit to the RCT receiving the highest quality rating, based within smoking cessation services, involving smokers motivated to quit and incorporating comprehensive face-to-face behavioural therapy. In accordance with this variation, the proportion of smokers quitting successfully differed markedly between trials. The generalisability of the RCT results across community, workplace and clinical contexts is unclear. It is likely that ENDS will be used differently by smokers who intend to quit and those who do not. Furthermore, the impact of any form of nicotine replacement is likely to differ according to whether or not it is used in conjunction with behavioural therapy and other support from smoking cessation services.^54^

The results of this systematic review are broadly consistent with the conclusions of earlier major reviews^13-17^ and more contemporary systematic reviews and meta-analyses,^19-21,55^ noting the overall paucity and generally uncertainty of the evidence. Of the three most recent systematic reviews and meta-analyses, two came to similar conclusions as ours and one – the most recent Cochrane Review – considered the evidence that ENDS was more efficacious for smoking cessation than ENNDS or NRT was moderate-certainty. However, this review included one study which did not have verified outcomes at six months,^56^ included some unpublished non-peer-reviewed data and gave overall higher quality ratings than ours. It also used fixed effects meta-analyses which assume a single underlying intervention effect and largely ignore heterogeneity.^57^ We chose *a priori* to use a random effects meta-analyses for our primary analyses, since we considered the included RCTs differed in terms of the nature of the intervention and populations being examined, including that devices and nicotine doses differed, participants were derived from different populations (e.g. motivated and unmotivated smokers, employees, general populations attending smoking cessation clinics and populations exposed to different tobacco control environments), background e-cigarette use prevalence varied and interventions were delivered differently, in ways likely to affect their effectiveness. Random-effects models also generally produced more conservative estimates but are less appropriate when there are few studies to summarise. In keeping with recommendations from the Cochrane Handbook,^58^ where there is uncertainty about which method is optimal, we also examined outcomes using fixed-effects meta-analyses, which showed results similar to the Cochrane review^21^ (Supplementary table S5). Our review is independent of the trials conducted to date, whereas three of the Cochrane review authors were authors of 3 of the 9 main trials included in the review and 2 of the 3 comparing ENDS and NRT. We and other recent independent systematic reviews are more cautious in our conclusions. For example, the Cochrane review considered there was moderate-certainty evidence, “limited by imprecision” that quit rates were higher in people randomised to nicotine versus non-nicotine e-cigarettes, in relation to a finding where the lower confidence bound was 1.00. Our findings are also consistent with the recent Irish Health Review Board’s independent network meta-analysis.^20^

Regardless of the analytic methods used and their interpretation, the fact remains that the results for each outcome are based on 3-4 studies, with methodological issues and limited numbers of participants and vary somewhat according to the type of analysis conducted; they are therefore not robust to the underlying analytic assumptions made. The limited reliable evidence on ENDS and smoking cessation is particularly stark when considered in relation to the current and potential scale of their use. It is also important to bear in mind the fact that most smokers quitting successfully do so unaided and that there are other smoking cessation tools that have a large evidence base demonstrating their efficacy, with no evidence of associated increases in the likelihood of tobacco smoking initiation among non-smokers.^59-61^

This review provides a comprehensive and up-to-date quantitative overview of evidence from RCTs on the efficacy of e-cigarettes as a smoking cessation tool. It includes only published studies with biochemically verified evidence of sustained smoking abstinence. It explicitly and quantitatively considers evidence independent of and with potential competing interests. This is the first review to our knowledge to examine the efficacy of e-cigarettes for nicotine cessation, finding limited evidence available. Nicotine cessation was not the primary or secondary outcome in any RCT and biochemical methods to validate nicotine cessation are still being developed.^62-64^ It includes only RCTs; while observational data provide useful evidence on some elements of e-cigarette use and their health impacts, smokers who do and do not use e-cigarettes differ in ways likely to affect their underlying propensity to quit, including in their commitment to quitting, health and health behaviours.

Tobacco smoking is one of the greatest risks to health ever faced – both for the population and the individual. Contemporary evidence indicates it is responsible for over 10% of deaths worldwide and up to one-half to two-thirds of long-term smokers will die from their habit.^65-68^ Tackling the tobacco epidemic requires both avoiding uptake of smoking and increasing quitting in current smokers. There is strong evidence that use of ENDS by (generally youth) never-smokers substantially increases uptake of smoking of combustible cigarettes.^13,55,69,70^ This review, and others, finds insufficient or weak evidence that ENDS helps smokers to quit, and we also note that the limited evidence of a potential effect relates largely to use in a therapeutic context. Hence, the current evidence indicates that e-cigarette policy and practice supportive of tobacco control would avoid ENDS exposure in never-smokers and would see better evidence regarding use as a smoking cessation aid. Internationally, the policy approach to ENDS is highly variable, ranging from widespread availability as consumer goods, with large-scale use including by young people, to prohibition.^71^ Policy options targeting cessation would need to include strong measures to avoid exposure – including diversion – to ENDS use by never-smokers; experiences with regulation of alcohol and combustible tobacco indicate that widespread availability combined with measures such as legal age limits, health warnings and other recommendations result in use by populations outside the target groups which is often extensive and harmful. There has been a tendency to confuse therapeutic and other types of ENDS use, with large-scale non-therapeutic use as a consumer good often promoted via tacit or overt claims of efficacy for smoking cessation.^72,73^

In conclusion, there is insufficient reliable evidence to conclude that nicotine-delivering e-cigarettes are a more effective smoking cessation aid than no intervention, placebo, or other NRT. The limited evidence to date is promising that they may be helpful, supporting the need for additional high-quality large-scale RCTs in the area. RCTs also show that nicotine-delivering e-cigarettes can result in prolonged exposure to nicotine through ongoing exclusive e-cigarette use or dual-use if the smoker does not quit.

## Data Availability

Supplementary material provided separately

## Acknowledgments and funding statement

This systematic review and meta-analysis was conducted as part of an independent program of work examining the health impacts of e-cigarettes, funded by the Australian Government Department of Health. Professor Banks is supported by a Principal Research Fellowship from the National Health and Medical Research Council of Australia (reference: 1136128). The authors have no conflicts of interest to declare.

## Notes

### Competing Interest Statement

The authors have declared no competing interest.

