## Supplementary for "Systematic review and meta-analysis of evidence on the efficacy of e-cigarette use for sustained smoking and nicotine cessation"

### Index

#### Supplementary Tables

Table S1: Additional details from randomised controlled trials of e-cigarettes and smoking cessation . 3

#### Table S1: Additional details from randomised controlled trials of e-cigarettes and smoking cessation

| Authors, year and setting | Blinding type | Population | Experimental intervention and number of participants randomised to each arm | Control intervention and number of participants randomised to control | Plan to quit | Sample size (enrolled/completed) | Statements regarding funding | Potential competing interests |
|---|---|---|---|---|---|---|---|---|
| Bullen et al., 2013[1]C*<br><br>New Zealand<br><br>Adults from the general community intending to quit, responding to media invitation | Single blinding | Adult smokers in New Zealand | Intervention 1 (n=289)<br>Electronic nicotine delivery system (ENDS), 16 mg nicotine from 1 week before until 12 weeks after quit day<br><br>Intervention 2 (n=73)<br>Electronic non-nicotine delivery system (ENNDS) from 1 week before until 12 weeks after quit day | Nicotine patches (n=295)<br>21 mg nicotine patch, one daily accessed via exchanging a voucher received in mail for patches at a community pharmacy | Yes | Intervention 1<br>289/241<br><br>Intervention 2<br>73/57<br><br>Control<br>295/215<br><br>Total<br>657/513 | Health Research Council of New Zealand. The e-cigarettes and cartridges were Elusion brand products provided by PGM International, New Zealand. | Yes |
| Caponnetto et al., 2013[2]*<br><br>Italy<br><br>Smokers not intending to quit were invited to try the 'Categoria' e-cigarette to reduce the risk of tobacco smoking | Double blinding | Adult smokers from Catania, Italy | Group A (n=100)<br>E-cigarette loaded with 7.2 mg for 12 weeks<br><br>Group B (n=100)<br>E-cigarette with 7.2 mg nicotine cartridge for 6 weeks and 5.4 mg nicotine cartridges for 6 weeks | Group C (n=100)<br>E-cigarettes with 12 week supply of non-nicotine cartridges | No | Intervention<br>Group A: 100/65<br>Group B: 100/63<br><br>Control<br>Group C=100/55<br><br>Total<br>300/183 | This research was supported by a grant-in-aid from Lega Italiana AntiFumo. The study sponsor had no involvement in the study design, collection, analysis, and interpretation of data, the writing of the manuscript or the decision to submit the manuscript for publication. RP and PC are currently funded by the University of Catania, Italy. The e- cigarette supplier had no involvement in the study design, collection, analysis, and interpretation of data, the writing of the manuscript or the decision to submit the manuscript for publication. The ''Categoria'' electronic cigarette kit and cartridges were provided free of charge by the local distributor, Arbi Group Srl, Italy. | Yes |

* Potential competing interest noted for study author(s)

| | | | | | | | | |
|---|---|---|---|---|---|---|---|---|
| Carpenter et al., 2017[3]<br><br>United States<br><br>Non-treatment seeking smokers from the community, recruited via media | Not stated | Adults smokers in the local community in a south eastern US urban area; approximately 30% non-white | Intervention 1 (n=25)<br>E-cigarette with 16 mg/mL nicotine<br><br>Intervention 2 (n=21)<br>E-cigarette with 24 mg/mL nicotine | No intervention (n=22) | Mixed | Intervention 1<br>25/19<br><br>Intervention 2<br>21/15<br><br>Control<br>22/16<br><br>Total<br>68/50 | Support was provided by NIH R21 DA037407 (to M.J. Carpenter), P01 CA200512 (to K.M. Cummings, M.J. Carpenter, and M.L. Goniewicz), UL1 TR001450, and P30 CA138313. M.L.Goniewicz's laboratory is supported via P30CA016056. B.W.Heckman is supported via K12 DA031794 and K23 DA041616. | No |
| Baldassarri et al. 2018[4]<br><br>United States<br><br>Hospital outpatient pulmonary and primary care clinics, Tobacco Treatment Service, and medical providers referrals | Double blinding | Treatment-seeking adult smokers from New Haven, Connecticut | Intervention (n=20)<br>E-cigarettes with 8 week supply of 24 mg/mL nicotine containing e-liquid, nicotine patch and counselling | Control (n=20)<br>E-cigarette with 8 week supply of 0 mg/mL nicotine containing e-liquid, nicotine patch and counselling | Yes | Intervention<br>20/unknown<br><br>Control<br>20/unknown<br><br>Total<br>40/unknown | Funding was provided by the Yale School of Medicine, Section of Pulmonary, Critical Care, and Sleep Medicine and the National Heart, Lung, and Blood Institute grant T32HL007778. | No |
| Halpern et al., 2018[5]*<br><br>United States<br><br>Employees and their spouses at 54 companies that used Vitality wellness programs | Not stated | Adult smokers who were employees or their spouses at 54 companies that used Vitality wellness programs across the United States | Intervention (n=1199)<br>NJOY e-cigarettes with up to 20 chambers of 1.0-1.5% nicotine content per week in participants' chosen flavours | Usual care (n=813)<br>Invitation to register for web-based smoking cessation, including information regarding the health benefits of smoking cessation, strategies to promote cessation, and the opportunity to register for the SmokeFreeTXT program of the National Cancer Institute | Mixed | Intervention<br>1199/253<br><br>Control<br>813/129<br><br>Total<br>2012/382 | Supported by a grant from the Vitality Institute to the University of Pennsylvania Center for Health Incentives and Behavioral Economics. | Yes |

* Potential competing interest noted for study author(s)

| | | | | | | | | |
|---|---|---|---|---|---|---|---|---|
| Hajek et al., 2019[6]<br><br>United Kingdom<br><br>Adults attending UK National Health Service stop-smoking services | Single blinding | Adult smokers from London | Intervention (n=438)<br>One 30mL bottle containing 18 mg/mL nicotine. Behavioural support including weekly one-on-one sessions with local clinicians | Nicotine-replacement (n=446)<br>Range of NRT products (patch, gum, lozenge, nasal spray, inhalator, mouth spray, mouth strip, and microtabs) and preferred product selected. Use of combinations was encouraged and participants were free to switch products. Behavioural support including weekly one-on-one sessions with local clinicians | Yes | Intervention<br>438/356<br><br>Control<br>446/342<br><br>Total<br>884/698 | Supported by the National Institute for Health Research (NIHR) Health Technology Assessment Programme (project number, 12/167/135) and by a grant (A16893) from the Cancer Research UK Prevention Trials Unit. | No |
| Lee et al., 2019[7]<br><br>Korea<br><br>Korean males from a motor company intending to quit | Single blinding | Male adult smokers employed at a motor company in Korea | Intervention (n=75)<br>E-cigarette containing 0.01 mg/mL nicotine for 12 weeks | Nicotine gum (n=75)<br>12 week supply of nicotine gum | Yes | Intervention<br>75/71 at 24 weeks<br><br>Control<br>75/61 at 24 weeks<br><br>Total<br>150/132 | None | No |
| Lucchiari et al. 2019[8]<br><br>Italy<br><br>COSMOS II lung cancer screening participants at the European Institute of Oncology (IEO) Hospital | Double blinding | Adult (≥55 years) chronic smokers participating in the COSMOS II program | Intervention 1 (n=70)<br>e-cigarette with 12 10mL liquid cartridges (8 mg/mL nicotine), telephone counselling<br><br>Intervention 2 (n=70)<br>e-cigarette with 12 10mL nicotine-free liquid cartridges, telephone counselling | Usual care (n=70)<br>Antismoking telephone counselling including phone interviews at weeks 1, 4, 8, and 12 | Yes | Intervention 1<br>70/52<br><br>Intervention 2<br>70/51<br><br>Control<br>70/52<br><br>Total<br>210/155 | Supported by Fondazione Umberto Veronesi (FUV). | No |

* Potential competing interest noted for study author(s)

| | | | | | | | | |
|---|---|---|---|---|---|---|---|---|
| Walker et al., 2019[9]*<br><br>New Zealand<br><br>Smokers from the community who were motivated to quit, recruited through media | Double blinding | Adult smokers in New Zealand | Intervention 1 (n=500)<br>ENDS (60:40 propylene glycol to vegetable glycerin ratio), a masked nicotine content of 0 mg/mL and a 21 mg, 24 h nicotine patch<br><br>Intervention 2 (n=499)<br>ENDS (60:40 propylene glycol to vegetable glycerin ratio), a masked nicotine content of 18 mg/mL and a 21 mg, 24 h nicotine patch | Nicotine patch only (n=125)<br>21 mg, 24 h nicotine patch | Yes | Intervention 1<br>499/337<br><br>Intervention 2<br>500/339<br><br>Control<br>125/63<br><br>Total<br>1124/739 | Health Research Council of New Zealand. | Yes |

* Potential competing interest noted for study author(s)

#### Table S2: Details of randomised controlled trials of e-cigarettes and smoking cessation, with data on nicotine use at follow up

| Authors, year, country and participants | Duration (treatment and follow-up) | Experimental intervention (n= randomised participants) | Control intervention (n= randomised participants) | Participants not using any nicotine at follow-up: ENDS, NRT or conventional cigarettes | Participants using NRT or ENDS at follow-up | Quitters using NRT or ENDS at follow-up | Non-quitters using NRT or ENDS at follow up |
|---|---|---|---|---|---|---|---|
| Bullen et al., 2013[1]*<br><br>New Zealand<br><br>Smokers from the general community intending to quit, responding to media invitation | Treatment<br>12 week supply received via courier or mailed voucher<br><br>Follow-up<br>1, 3, 6 months via telephone and 6 month laboratory visit for those reporting cessation | Intervention 1 (n=289)<br>Electronic nicotine delivery system (ENDS), 16 mg nicotine from 1 week before until 12 weeks after quit day<br><br>Intervention 2 (n=73)<br>Electronic non-nicotine delivery system (ENNDS) from 1 week before until 12 weeks after quit day | Nicotine patches (n=295)<br>21 mg nicotine patch, one daily accessed via exchanging a voucher received in mail for patches at a community pharmacy | ENDS: 4.5% (12/289)<br>Patches: Not stated<br>ENNDS: 4.1% (3/73)* | **Adherence at 6 months**<br>ENDS: 24.6% (71/289)<br>Patches: 5.8% (17/295)<br><br>**Relative Risk (95% CI)****<br>ENDS vs patches<br>4.26 (2.58-7.06)<br><br>Reported in the paper per protocol<br>ENDS: 29% (71/241)<br>Patches: 8% (12/215) | ENDS: 38% (8/21)<br>Patches: Not stated | ENDS: 29% (63/220)<br>Patches: Not stated<br><br>(NB: Unclear whether ENDS or ENNDS) |
| Caponnetto et al., 2013[2]*<br><br>Italy<br><br>Smokers not intending to quit invited via newspaper advertisements to “try e-cigarettes to reduce the | Treatment<br>12 weeks dispensed at baseline visit held at smoking cessation clinic<br><br>Follow-up<br>2, 4, 6, 8, 10, 12, 24 and 52 | Group A (n=100)<br>E-cigarette loaded with 7.2 mg for 12 weeks<br><br>Group B (n=100)<br>E-cigarette with 7.2 mg nicotine cartridge for 6 weeks and 5.4 mg nicotine cartridges for 6 weeks | Group C (n=100)<br>E-cigarettes with 12 week supply of non-nicotine cartridges | Not stated | Not stated | Group A, B & C:<br>26.9% (7/26)<br><br>(NB: Unclear whether ENDS or ENNDS) | Not stated |

| | | | | | | | |
|---|---|---|---|---|---|---|---|
| risk of tobacco smoking." | week visits to study clinic | | | | | | |
| Carpenter et al., 2017[3]<br><br>United States<br><br>Non-treatment seeking smokers from the community, recruited via media | Treatment<br>3 weeks, laboratory visits at 2, 3 and 4 weeks<br><br>Follow-up<br>Laboratory visits at 8, 12, and 16 weeks | Intervention 1 (n=25)<br>E-cigarette with 16 mg/mL nicotine<br><br>Intervention 2 (n=21)<br>E-cigarette with 24 mg/mL nicotine | No intervention (n=22) | Not stated | **ENDS use at week 16**<br>Intervention 1<br>32% (8/25)<br>Intervention 2<br>60% (13/21)<br>Control<br>13% (3/22) | Not stated | Not stated |
| Baldassarri et al. 2018[4]<br><br>United States<br><br>Motivated smoking patients from hospital outpatient pulmonary and primary care clinics, tobacco treatment service, and medical provider referrals | Treatment<br>8 weeks, laboratory visits at 2, 4, 6, and 8 weeks<br><br>Follow-up<br>Laboratory visit at 24 weeks | Intervention (n=20)<br>E-cigarettes with 8 week supply of 24 mg/mL nicotine containing e-liquid, nicotine patch and counselling | Control (n=20)<br>E-cigarette with 8 week supply of 0 mg/ml nicotine containing e-liquid, nicotine patch and counselling | ENNDS + patch: 5% (1/20)<br>ENDS + patch: 10% (2/20)<br><br>**Relative Risk (95% CI)****<br>ENDS + patch vs ENNDS + patch<br>2.00 (0.20-20.33) | Not stated | ENNDS + patch: 50% (1/2)<br>ENDS + patch: 50% (2/4)<br><br>**Relative Risk (95% CI)****<br>ENDS + patch vs ENNDS + patch<br>1.00 (0.18-5.46) | Not stated |
| Hajek et al., 2019[6]<br><br>United Kingdom<br><br>Adults attending UK National Health Service stop-smoking | Treatment<br>12 weeks, trial visit at enrolment and week 4<br><br>Follow-up | Intervention (n=438)<br>One 30mL bottle containing 18 mg/mL nicotine. Behavioural support including weekly one-on-one sessions with local clinicians | Nicotine-replacement (n=446)<br>Range of NRT products (patch, gum, lozenge, nasal spray, inhalator, mouth spray, mouth strip, and microtabs) and preferred product | ENDS: 3.65% (16/438)<br>NRT: 8.97% (40/446)<br><br>**Relative Risk (95% CI)****<br>ENDS vs other NRT<br>0.41 (0.23-0.72) | **Adherence at 52 weeks**<br>ENDS: 39.5% (173/438)<br>NRT: 4.3% (19/446)<br><br>**Relative Risk (95% CI)****<br>ENDS vs other NRT<br>9.27 (5.88-14.61) | ENDS: 80% (63/79)<br>NRT: 9% (4/44)<br><br>**Relative Risk (95% CI)****<br>ENDS vs other NRT<br>8.77 (3.42-22.48) | ENDS: 30.6% (110/359)<br>NRT: 3.7% (15/402)<br><br>**Relative Risk (95% CI)****<br>ENDS vs other NRT<br>8.21 (4.88-13.82) |

| | | | | | | | |
|---|---|---|---|---|---|---|---|
| services | 52 weeks, phone call at 26 and 52 weeks and trial visit at 52 weeks | | selected. Use of combinations was encouraged and participants were free to switch products. Behavioural support including weekly one-on-one sessions with local clinicians | | | | |
| Walker et al., 2019[9]*<br><br>New Zealand<br><br>Smokers from the community who were motivated to quit, recruited through media | Treatment<br>12 weeks, 14 week supply delivered by courier<br><br>Follow-up<br>6 months after quit date, phone call at 1, 3, and 6 months, clinic visit at 6 months for those reporting cessation | Intervention 1 (n=500)<br>ENDS (60:40 propylene glycol to vegetable glycerin ratio), a masked nicotine content of 0 mg/mL and a 21 mg, 24 h nicotine patch<br><br>Intervention 2 (n=499)<br>ENDS (60:40 propylene glycol to vegetable glycerin ratio), a masked nicotine content of 18 mg/mL and a 21 mg, 24 h nicotine patch | Nicotine patch only (n=125)<br>21 mg, 24 h nicotine patch | Not stated | **Adherence at 6 months**<br>Control:<br>21/52 (40%)<br>Intervention 1<br>Both: 41/308 (13%)<br>ENNDS only: 111/308 (36%)<br>Patch only 88/308 (29%)<br>Intervention 2<br>Both: 36/317 (11%)<br>ENDS only: 143/317 (45%)<br>Patch only: 70/317 (22%)<br><br>**Relative Risk (95% CI)****<br>Patch + ENDS vs Patch only<br>1.53 (1.05-2.22)<br>Patch + ENNDS vs Patch only<br>1.52 (1.05-2.21)<br>Patch + ENDS vs Patch + ENNDS<br>1.00 (0.88-1.15) | Not stated | Not stated |

#### Table S3: Risk of bias assessment of randomised controlled trials of e-cigarettes for smoking cessation

| Study | Randomisation process | Deviations from intended interventions | Missing outcome data | Measurement of the outcome | Risk of bias in selection of the reported result | Overall judgment |
|---|---|---|---|---|---|---|
| Bullen et al. 2013[1]* | Low | Some concerns | Low | Low | Low | Some concerns |
| Caponnetto et al 2013[2]* | Low | Some concerns | High | Low | Some concerns | High |
| Carpenter et al. 2017[3] | Some concerns | Some concerns | Low | High | Some concerns | High |
| Baldassarri et al. 2018[4] | Low | Low | High | Low | Some concerns | High |
| Halpern et al., 2018[5]* | Some concerns | Some concerns | High | Low | Low | High |
| Hajek et al., 2019[6] | Low | Low | Low | Low | Low | Low |
| Lee et al., 2019[7] | Low | Some concerns | High | Low | Some concerns | High |
| Lucchiari et al. 2019[8] | Low | Some concerns | Some concerns | Low | Low | Some concerns |
| Walker et al., 2019[9]* | Low | Low | High | Low | Low | High |

#### Table S4: GRADE assessment of randomised controlled trials of e-cigarettes for smoking cessation

| **Outcome** | **Risk of bias** | **Indirectness** | **Inconsistency** | **Imprecision** | **Publication bias** | **Quality of the evidence** |
|---|---|---|---|---|---|---|
| Combustible tobacco cigarette smoking cessation | Serious concern[1] | No concern | No concern | Serious concern[2] | Undetected | Low |

[1]Downgraded based on the overall risk of bias assessment from the ROB2 tool.

[2]Downgraded due to small number of events across all studies and presence of wide confidence intervals.

#### Table S5. Sensitivity analysis: meta-analysis of randomised controlled trials of e-cigarettes for smoking cessation including random and fixed effects models

| Study | Treatment / Follow up duration (weeks) | Outcome | | Risk ratio (95% CI) | Random effects | | Fixed effects | |
|---|---|---|---|---|---|---|---|---|
| | | Intervention % (Events/Total) | Control % (Events/Total) | | % weight | Risk ratio (95% CI) | % weight | Risk ratio (95% CI) |
| A. Nicotine-delivering e-cigarettes versus no intervention or usual care | | | | | | | | |
| Carpenter et al. 2017^ | 3 / 16 | 6.5% (3/46) | 4.5% (1/22) | 1.43 (0.16-13.02) | 12.21 | 1.96 (0.91-4.23) | 15.12 | 2.08 (0.96-4.48) |
| Halpern et al. 2018^# | 26 / 26 | 1.0% (4/1199) | 0.0% (1/813) | 6.11 (0.33-113.24) | 6.96 | | 6.66 | |
| Lucchiari et al. 2019^ | 12 / 26 | 18.5% (13/70) | 10% (7/70) | 1.86 (0.78-4.38) | 80.83 | | 78.22 | |
| B. Nicotine-delivering e-cigarettes versus non-nicotine-e-cigarettes | | | | | | | | |
| Bullen 2013* | 12 / 26 | 7.3% (21/289) | 4.1% (3/73) | 1.77 (0.54-5.77) | 20.81 | 1.61 (0.93-2.78) | 22.68 | 1.71 (1.00-2.92) |
| Caponetto 2013* | 12 / 52 | 11% (22/200) | 4% (4/100) | 2.75 (0.97-7.76) | 26.8 | | 25.25 | |
| Lucchiari et al. 2019^ | 12 / 26 | 18.5% (13/70) | 15.7% (11/70) | 1.18 (0.57-2.46) | 52.39 | | 52.07 | |
| C. Nicotine-delivering e-cigarettes versus other nicotine-replacement therapy | | | | | | | | |
| Bullen et al. 2013* | 12 / 26 | 7.3% (21/289) | 5.8% (17/295) | 1.26 (0.68-2.34) | 28.62 | 1.25 (0.75-2.10) | 20.41 | 1.43 (1.10-1.86) |
| Hajek et al. 2019 | 12 / 52 | 18.0% (79/438) | 9.9% (44/446) | 1.83 (1.30-2.58) | 39.91 | | 52.9 | |
| Lee et al. 2019^ | 12 / 24 | 22.7% (17/75) | 29.3% (22/75) | 0.77 (0.45-1.33) | 31.47 | | 26.69 | |

* Potential competing interests have been noted

^ RRs are calculated from number of events or percentages reported in the published study

### RR is undefined due to zero events in the control group. RR estimated by applying the continuity correction (adding 0.5 to each cell of the 2x2 table)

#### Figure S1: Sensitivity analysis: verified smoking cessation in smokers randomised to nicotine-delivering e-cigarettes versus no intervention or usual care in studies with no reported potential competing interests

| Study | Treatment/Follow up duration (weeks) | Intervention % (Events/Total) | Control % (Events/Total) | Risk Ratio with 95% CI |
|---|---|---|---|---|
| Carpenter et al. 2017^ | 3/16 | 6.5% (3/46) | 4.5% (1/22) | 1.43 [ 0.16, 13.02] |
| Lucchiari et al. 2019^ | 12/26 | 18.5% (13/70) | 10.0% (7/70) | 1.86 [ 0.79, 4.38] |
| **Overall** | | | | 1.80 [ 0.81, 3.99] |

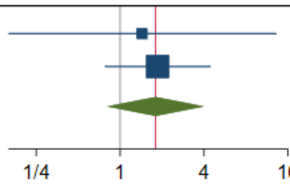

^ RRs are calculated from number of events or percentages reported in the published study

Total cessation events: 16/116 in intervention group, 8/92 in control group

Heterogeneity: $Tau^2$=0.00; $Chi^2$= 0.05, df=1,, p = 0.83; $I^2$ =0.0%; Test for overall effect: Z=1.44, p=0.15

#### Figure S2: Sensitivity analysis: verified smoking cessation in smokers randomised to nicotine-delivering e-cigarettes versus other nicotine-replacement therapy in studies with no reported potential competing interests

| Study | Treatment/Follow up duration (weeks) | Intervention % (Events/Total) | Control % (Events/Total) | Risk Ratio with 95% CI |
|---|---|---|---|---|
| Hajek et al. 2019 | 12/52 | 18.0% (79/438) | 9.9% (44/446) | 1.83 [ 1.30, 2.58] |
| Lee et al. 2019^ | 12/24 | 21.3% (16/75) | 28.0% (21/75) | 0.76 [ 0.43, 1.34] |
| **Overall** | | | | 1.22 [ 0.52, 2.86] |

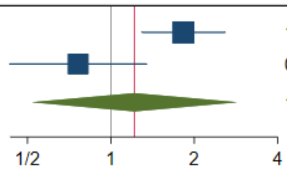

^ RRs are calculated from number of events or percentages reported in the published study

Total cessation events: 95/513 in intervention group, 65/521 in control group

Heterogeneity: $Tau^2$=0.00; $Chi^2$= 6.70, df=1, p = 0.01; $I^2$ =85.1%; Test for overall effect: Z=0.45, p=0.65

#### Figure S3: Sensitivity analysis: verified smoking cessation in smokers randomised to nicotine-delivering e-cigarettes versus no intervention or usual care at 6-month follow-up

| Study | Treatment/Follow up duration (weeks) | Intervention % (Events/Total) | Control % (Events/Total) | Risk Ratio with 95% CI |
|---|---|---|---|---|
| Halpern et al. 2018^* | 26/26 | 1.0% (12/1199) | 0.1% (1/813) | 8.14 [ 1.06, 62.45] |
| Lucchiari et al. 2019^ | 12/26 | 18.5% (13/70) | 10.0% (7/70) | 1.86 [ 0.79, 4.38] |
| **Overall** | | | | 2.88 [ 0.77, 10.79] |

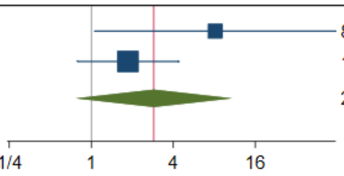

Total cessation events: 20/1315 in intervention group, 8/905 in control group

Heterogeneity: Tau$^2$=0.46; Chi$^2$= 1.72, df=1, p = 0.19; I$^2$ =41.7%; Test for overall effect: Z=1.57, p=0.12

#### Figure S4: Sensitivity analysis: verified smoking cessation in smokers randomised to nicotine-delivering e-cigarettes versus non-nicotine delivering e-cigarettes at 6-month follow-up

| Study | Treatment/Follow up duration (weeks) | Intervention % (Events/Total) | Control % (Events/Total) | Risk Ratio with 95% CI |
|---|---|---|---|---|
| Bullen et al. 2013* | 21/26 | 7.3% (21/289) | 41% (3/73) | 1.77 [ 0.54, 5.77] |
| Caponetto et al. 2013* | 12/24 | 11.0% (22/200) | 5.0% (5/100) | 2.20 [ 0.86, 5.64] |
| Lucchiari et al. 2019^ | 12/26 | 18.5% (13/70) | 15.7% (11/70) | 1.18 [ 0.57, 2.46] |
| **Overall** | | | | 1.54 [ 0.92, 2.59] |

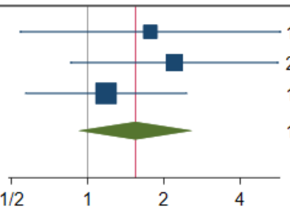

Total cessation events: 20/1315 in intervention group, 8/905 in control group

Heterogeneity: Tau$^2$=0.00; Chi$^2$= 1.11, df=2, p = 0.57; I$^2$ =0.0%; Test for overall effect: Z=1.64, p=0.10

#### Figure S5: Sensitivity analysis: verified smoking cessation in smokers randomised to nicotine-delivering e-cigarettes versus nicotine replacement therapy at 6-month follow-up

| Study | Treatment/Follow up duration (weeks) | Intervention % (Events/Total) | Control % (Events/Total) | Risk Ratio with 95% CI |
|---|---|---|---|---|
| Bullen et al. 2013* | 12/26 | 7.3% (21/295) | 5.8% (17/295) | 1.26 [ 0.68, 2.34] |
| Hajek et al. 2019 | 12/26 | 35.4% (155/438) | 25.1% (112/446) | 1.41 [ 1.15, 1.73] |
| Lee et al. 2019^ | 12/24 | 21.3% (16/75) | 28.0% (21/75) | 0.76 [ 0.43, 1.34] |
| **Overall** | | | | 1.18 [ 0.82, 1.70] |

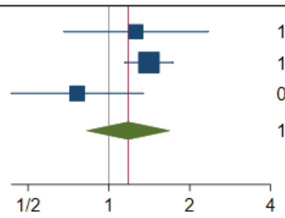

Total cessation events: 20/1315 in intervention group, 8/905 in control group

Heterogeneity: Tau$^2$=0.00; Chi$^2$= 4.02, df=2, p = 0.13; I$^2$ =50.5%; Test for overall effect: Z=0.89, p=0.37

### Appendix 1: Additional methodological details for systematic review of e-cigarettes and smoking cessation

#### Search Strategy

##### MEDLINE search terms:

1. Smoker.mp
2. Smokers.mp
3. Ex-Smokers.mp
4. Ex-Smokers.mp
5. Exp Smokers/
6. Exp Ex-smokers/
7. 1 or 2 or 3 or 4 or 5 or 6
8. E-cigarette.mp
9. E-cigarettes.mp
10. “electronic cigarette”.mp
11. “electronic cigarettes”.mp
12. “electronic nicotine de*”.mp
13. “electronic nicotine delivery system”.mp
14. Vape.mp
15. Vaping.mp
16. Vapo*.mp
17. E-liquid.mp
18. E-hookah.mp
19. “Electronic inhalant device”.mp
20. Exp “Electronic nicotine delivery systems”/
21. 8 or 9 or 10 or 11 or 12 or 13 or 14 or 15 or 16 or 17 or 18 or 19 or 20
22. “Smoking cessation”.mp
23. Cessation.mp
24. Quit.mp
25. Abstinence.mp
26. Exp “smoking cessation”/
27. Exp “tobacco use cessation devices”/
28. Exp “smoking cessation agents”/
29. 22 or 23 or 24 or 25 or 26 or 27 or 28
30. 7 and 21 and 29
31. Limit 30 to randomized controlled trials

Results: 96

##### PsycINFO search terms:

1. Smoker.mp
2. Smokers.mp
3. Ex-Smokers.mp
4. Ex-Smokers.mp
5. Smokers.mh
6. Ex-smokers.mh
7. 1 or 2 or 3 or 4 or 5 or 6
8. E-cigarette.mp
9. E-cigarettes.mp
10. “electronic cigarette”.mp
11. “electronic cigarettes”.mp
12. “electronic nicotine de*”.mp
13. “electronic nicotine delivery system”.mp
14. Vape.mp

15. Vaping.mp
16. Vapo*.mp
17. E-liquid.mp
18. E-hookah.mp
19. “Electronic inhalant device”.mp
20. “Electronic nicotine delivery systems”.mh
21. 8 or 9 or 10 or 11 or 12 or 13 or 14 or 15 or 16 or 17 or 18 or 19 or 20
22. “Smoking cessation”.mp
23. Cessation.mp
24. Quit.mp
25. Abstinence.mp
26. “Smoking cessation”.mh
27. “Tobacco use cessation devices”.mh
28. “Smoking cessation agents”.mh
29. 22 or 23 or 24 or 25 or 26 or 27 or 28
30. 7 and 21 and 29
31. Limit 30 to “0300 clinical trial”

Results: 13

#### PubMed search terms:

1. ((("smoking cessation" OR Cessation OR quit OR Abstinence OR "smoking cessation" [MeSH Terms] OR "tobacco use cessation devices"[MeSH Terms] OR "smoking cessation agents"[MeSH Terms]) AND (E-cigarette OR E-cigarettes OR "Electronic cigarette" OR "Electronic cigarettes" OR "Electronic nicotine de*" OR "Electronic nicotine delivery system" OR Vape OR Vaping OR E-liquid OR Vapo* OR E-hookah OR "Electronic inhalant device" OR "Electronic nicotine delivery systems"[MeSH Terms]) AND (Smoker OR Smokers OR Ex-smoker OR Ex smokers OR Smokers[MeSH Terms] OR Exsmokers[MeSH Terms]))) AND Randomized Controlled Trial[ptyp]

Results: 87

#### Scopus search terms:

1. TITLE-ABS-KEY (("smoking cessation" OR Cessation OR quit OR Abstinence OR "tobacco use cessation devices" OR "smoking cessation agents") AND (E-cigarette OR E-cigarettes OR "Electronic cigarette" OR "Electronic cigarettes" OR "Electronic nicotine de*" OR "Electronic nicotine delivery system" OR Vape OR Vaping OR E-liquid OR Vapo* OR E-hookah OR "Electronic inhalant device") AND (Smoker OR Smokers OR Ex-smoker OR Ex-smokers) AND (LIMIT-TO (DOCTYPE, “ar”)))

Results: 3,759

#### Web of Science search terms:

2. TS=("smoking cessation" OR Cessation OR quit OR Abstinence) AND TS=(E-cigarette OR E cigarettes OR "Electronic cigarette" OR "Electronic cigarettes" OR "Electronic nicotine de*" OR "Electronic nicotine delivery system" OR Vape OR Vaping OR E-liquid OR Vapo* OR E-hookah OR "Electronic inhalant device") AND TS=(Smoker OR Smokers OR Ex-smoker OR Ex-smokers)) AND DOCUMENT TYPES: (Article)
   Indexes=SCI-EXPANDED, SSCI, A&HCI, CPCI-S, CPCI-SSH, ESCI, CCR-EXPANDED, IC Timespan=All years

Results: 930

#### Cochrane search terms:

1. (Smoker):ti,ab,kw OR (Smokers):ti,ab,kw OR (Exsmoker): ti,ab,kw OR (Ex-smokers):ti,ab,kw
2. MeSH descriptor: [Smokers] explode all trees
3. MeSH descriptor: [Ex-Smokers] explode all trees
4. #1 OR #2 OR #3
5. E-cigarette OR E-cigarettes OR "Electronic cigarette" OR "Electronic cigarettes" OR "Electronic nicotine de*" OR "Electronic nicotine delivery system" OR Vape OR Vaping OR E liquid OR Vapo* OR E-hookah OR "Electronic inhalant device"

6. MeSH descriptor: [Electronic Nicotine Delivery Systems] explode all trees
7. #5 OR #6
8. "smoking cessation" OR Cessation OR quit OR Abstinence
9. MeSH descriptor: [Smoking Cessation] explode all trees
10. MeSH descriptor: [Tobacco Use Cessation Devices] explode all trees
11. MeSH descriptor: [Smoking Cessation Agents] explode all trees
12. #8 OR #9 OR #10 OR #11
13. #4 AND #7 AND #12
14. #13 in trials

Results: 2

#### Inclusion and Exclusion Criteria

**Inclusion criteria:**

Study designs: Published, peer-reviewed randomised control trials

Population: Current tobacco smokers, humans, any age, no limit on smoking status (duration, cigarettes per day etc.), smokers motivated or unmotivated to quit

Intervention: Nicotine-containing or non-nicotine-containing e-cigarettes or e-liquids

Comparison: No e-cigarettes, placebo
Standard smoking cessation treatment/aids such as Nicotine Replacement Therapies (e.g. patch, gum, inhalers), behavioural and/or pharmacological cessation aids (e.g. bupropion & varenicline), and combination of e-cigarettes and treatments
Any other treatments or aids intended to assist with cessation.

Outcome: Primary or secondary outcome variable is combustible tobacco smoking cessation.
RCT contains outcome data on cessation of nicotine exposure in any form and cessation of non-nicotine containing e-cigarettes.
Abstinence must be biochemically verified at a minimum 4 month follow up

Timing: All years

Setting: Any country

Language: Articles reported in English.

**Exclusion criteria:**

Study designs: Systematic reviews and meta-analyses, non-systematic reviews – literature reviews, non-randomised clinical trial, intervention trial with no comparator (e.g. before and after study), qualitative studies, prospective cohort studies / cross over trials, retrospective cohort studies, cross-sectional studies, case-control studies, case studies, grey literature, conference abstracts, letters, editorials, correspondence, opinion pieces, government reports, position statements

Population: In vitro studies or animal studies

Intervention: Heat-not-burn and tobacco containing products

Outcome: Studies where smoking, or nicotine, cessation is not the primary or secondary outcome variable.

Timing: No exclusion criteria.

Setting: No exclusion criteria.

Language: Articles not published or translated to English.

Other: Duplicated data, unavailable full text.

#### Data Extraction

Two authors of this review independently extracted data from the included RCTs using a pre-specified data extraction template. Available data on cessation of nicotine in any form (e.g. combustible tobacco, ENDS, other NRT); use of NRT, including ENDS, or ENNDS, among all participants; use of NRT, including e-cigarettes, or non-nicotine e-cigarettes, among quitters; and use of NRT, including e-cigarettes, or non-nicotine e-cigarettes, among those who do not quit, were extracted.
